# RBC storage duration does not affect biochemical recurrence after radical prostatectomy

**DOI:** 10.64898/2026.01.05.26343475

**Authors:** Michael Obuobi, Michael Asante Ofosu, Berlinda Nyantakyi, Saviour Ntow, Abigail Amoah, George Ekow Quaye

## Abstract

The relationship between red blood cell (RBC) storage duration and cancer recurrence remains controversial, with the ”storage lesion” potentially amplifying transfusion-related immunomodulation effects. This retrospective cohort study examined whether RBC storage duration is independently associated with biochemical recurrence following radical prostatectomy for prostate cancer. We analyzed 316 men who underwent radical prostatectomy with perioperative allogeneic RBC transfusion at Cleveland Clinic (1998-2007). Patients were stratified by RBC storage duration: younger (≤13 days, n=106), middle (13-18 days, n=103), and older (≥18 days, n=107). Primary outcome was biochemical recurrence (PSA ≥0.4 ng/mL). We employed Kaplan-Meier survival analysis, Cox proportional hazards regression, and multivariable logistic regression with comprehensive diagnostics. Missing data (10.8%) were handled using multiple imputation by chained equations (MICE). Over median follow-up of 36.2 months, 54 patients (17.1%) experienced biochemical recurrence. RBC storage groups demonstrated exceptional baseline balance (all p>0.05). Kaplan-Meier analysis showed virtually identical survival curves (log-rank p=0.98). In fully adjusted Cox regression, neither middle (HR=1.01, 95% CI: 0.47-2.18, p=0.987) nor older (HR=0.82, 95% CI: 0.38-1.76, p=0.569) RBC storage was associated with recurrence. Logistic regression yielded consistent results (middle: OR=0.85, p=0.692; older: OR=0.93, p=0.854). Effect estimates remained stable across progressive covariate adjustment. In contrast, tumor biology factors demonstrated powerful associations: Gleason 8-10 (OR=5.72, p=0.001), log(PSA) (OR=2.24, p=0.007), and organ confinement (OR=0.48, p=0.047). Model discrimination was excellent (C-index=0.798, AUC=0.805), driven entirely by tumor characteristics. RBC storage alone provided zero discriminative ability (AUC=0.511). RBC storage duration (10-25 days) is not associated with biochemical recurrence after radical prostatectomy. This null finding is consistent across three complementary analytical methods. Tumor biology dominates prognosis with effect sizes far exceeding any potential transfusion effects. Current blood banking practices appear oncologically safe within typical storage windows.

## Introduction

Prostate cancer remains the most common non-cutaneous malignancy in American men, with an estimated 288,300 new cases projected for 2023 [1]. Radical prostatectomy serves as definitive surgical treatment for localized disease, achieving excellent oncologic outcomes with 10-year cancer-specific survival rates exceeding 95% for low-risk disease [2]. Despite advances in minimally invasive techniques, perioperative blood transfusions remain necessary in 5-30% of cases, depending on surgical approach, surgeon experience, and patient characteristics [3]. Beyond immediate transfusion risks, mounting evidence suggests potential long-term effects on cancer outcomes through transfusion-related immunomodulation (TRIM), which encompasses immunosuppressive effects including reduced T-lymphocyte proliferation, altered cytokine profiles, and impaired natural killer cell function that may persist for weeks to months [4, 5]. These effects could theoretically facilitate tumor cell survival during the critical perioperative period when circulating tumor cells are released through surgical manipulation [6].

The ”storage lesion” refers to progressive biochemical, biomechanical, and immunologic changes occurring in refrigerated red blood cells during storage at 1-6°C for up to 42 days [7, 8]. These time-dependent changes include membrane vesiculation, ATP depletion (50-70% by 35 days), complete 2,3-DPG loss by 14-21 days, accumulation of pro-inflammatory cytokines and bioactive lipids, and rising extracellular potassium [9]. The hypothesis suggests older stored blood may confer greater immunosuppressive effects and worse clinical outcomes compared to fresher blood, with changes accelerating significantly after 2-3 weeks of storage [8].

While animal studies demonstrated that transfusion of blood stored >9 days significantly increased experimental lung metastases through NK cell suppression [10], clinical evidence remains inconsistent. In prostate cancer, the seminal Cleveland Clinic analysis found no association between RBC storage duration and biochemical recurrence (p=0.82) [11], though this employed relatively basic statistical methods. Three landmark randomized trials (ABLE, INFORM, RECESS) comparing fresh versus standard-issue blood found no mortality benefit in general populations [12–14], but specifically excluded cancer patients and did not examine oncologic outcomes. Critical knowledge gaps persist: limited analytical rigor in observational cancer studies, single analytical frameworks, inadequate model diagnostics, incomplete missing data handling, and insufficient exploration of effect modification by disease severity.

We conducted this comprehensive reanalysis to determine whether RBC storage duration is independently associated with biochemical recurrence following radical prostatectomy, using systematic statistical methods with thorough covariate adjustment and cross-validation across multiple analytical frameworks. Secondary objectives include comparing recurrence-free survival rates across storage groups using Kaplan-Meier analysis, quantifying independent RBC storage effects using Cox and logistic regression, identifying dominant prognostic factors, and evaluating potential effect modification by disease severity. We hypothesized that RBC storage duration would not demonstrate independent association with biochemical recurrence after multivariable adjustment, based on prior null findings, large trial evidence, and theoretical considerations including tumor biology dominance and transient immunosuppression effects.

## Materials and methods

### Study Design and Data Source

This retrospective cohort study utilized the BloodStorageProstate df dataset from the publicly available medicaldata R package [21], originally published by Cata et al. [11]. The dataset includes 316 men who underwent radical prostatectomy with perioperative allogeneic RBC transfusion at Cleveland Clinic (1998-2007), with data obtained from prospectively maintained registries electronically linked to the institutional blood bank database. The dataset was de-identified and made publicly available for research purposes. Data were accessed using R version 4.0.5.

The dataset contains comprehensive information on patient demographics, disease characteristics, pathologic features, transfusion details, and biochemical recurrence outcomes. All 316 patients were included and categorized into three RBC storage groups: younger (≤13 days, n=106, 33.5%), middle (13-18 days, n=103, 32.6%), and older (≥18 days, n=107, 33.9%). All patients had histologically confirmed prostate adenocarcinoma and at least one post-operative PSA measurement for biochemical recurrence assessment.

**Fig 1.**
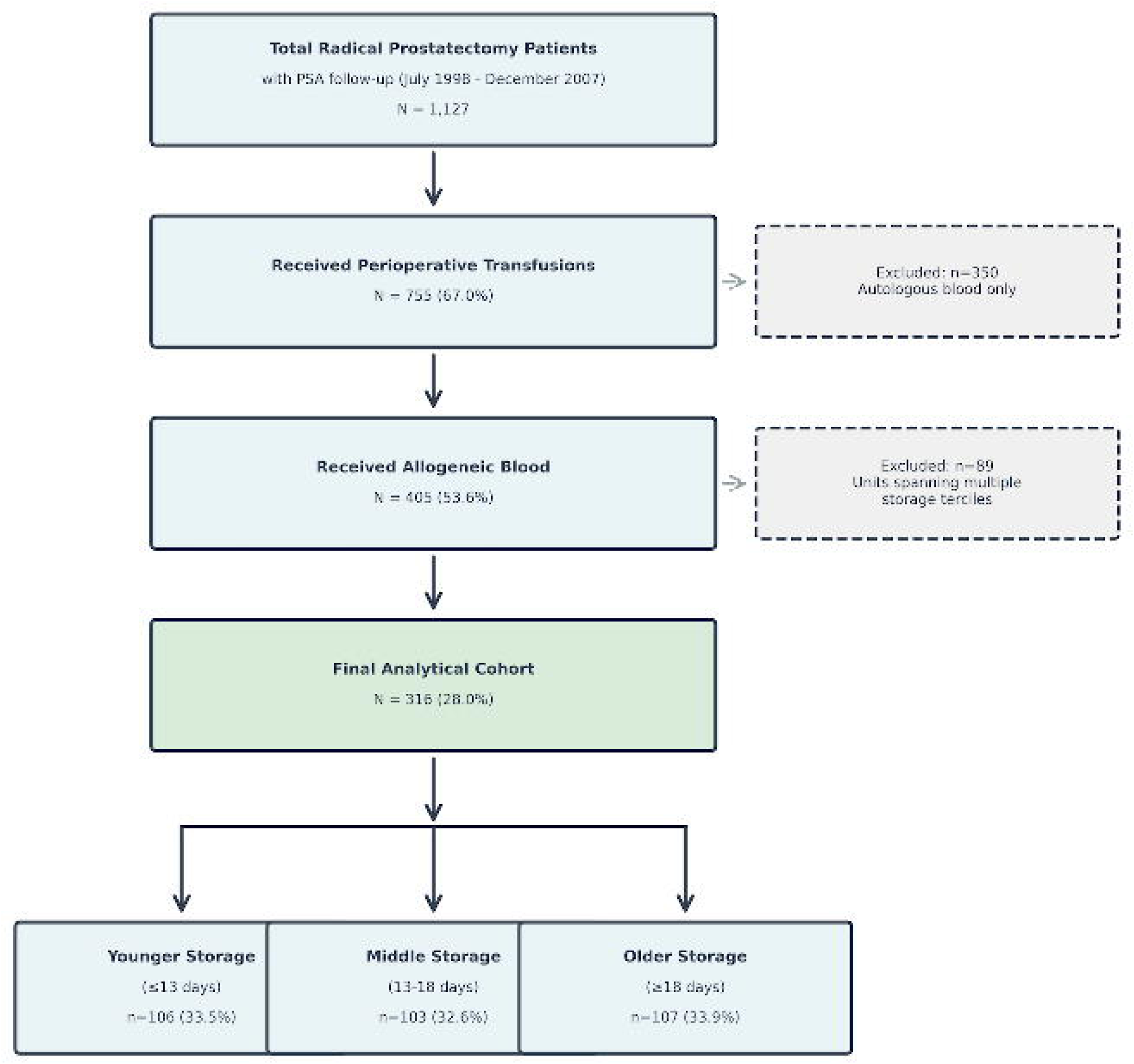
Study flow diagram showing patient selection and RBC storage group allocation. From 1,127 men who underwent radical prostatectomy with PSA follow-up (July 1998 - December 2007), 755 (67.0%) received perioperative transfusions. After excluding 350 patients who received autologous blood only and 89 with units spanning multiple storage terciles, the final analytical cohort comprised 316 patients (28.0%) stratified into three RBC storage terciles: younger storage (≤13 days, n=106, 33.5%), middle storage (13-18 days, n=103, 32.6%), and older storage (≥18 days, n=107, 33.9%).

### Exposure and outcome definitions

The primary exposure was RBC storage duration, calculated as the number of days from blood collection to transfusion. For patients receiving multiple units (median 2, range 1-19), the median storage duration was used. Patients were categorized into terciles: younger storage (≤13 days, n=106, 33.5%), middle storage (13-18 days, n=103, 32.6%), and older storage (≥18 days, n=107, 33.9%). Observed storage ranged from 10-25 days (median 15 days, IQR 12-18 days). The primary outcome was biochemical recurrence, defined as PSA ≥0.4 ng/mL on two consecutive measurements at least one month apart [15]. For survival analyses, time-to-event was calculated from surgery date to first PSA ≥0.4 ng/mL, with censoring at last PSA measurement or death. For logistic regression, biochemical recurrence was analyzed as a binary outcome (yes/no).

### Covariates

Potential confounders and prognostic factors were identified a priori based on established prostate cancer prognostic literature and biological plausibility for confounding the RBC-recurrence relationship. Covariates included three categories:

#### Demographics

Age at surgery (continuous, years), race (binary: African American vs. non-African American), and family history of prostate cancer (binary: yes vs. no, defined as first-degree relative with prostate cancer diagnosis).

#### Pre-treatment disease characteristics

Preoperative PSA (continuous, ng/mL, log-transformed for regression modeling due to right-skewed distribution), biopsy Gleason score (ordinal categorical: 0-6 [low grade/well-differentiated] vs. 7 [intermediate grade/moderately differentiated] vs. 8-10 [high grade/poorly differentiated]), and clinical T stage (binary: T1-T2a [localized, non-palpable or unilateral palpable] vs. T2b-T3 [bilateral palpable or locally advanced]).

#### Pathologic features

Pathologic organ confinement (binary: yes [pT2, organ-confined] vs. no [pT3, extraprostatic extension, seminal vesicle invasion, or bladder neck invasion]) and bladder neck margin status (binary: positive [cancer cells at inked surgical margin] vs. negative).

All covariates were measured at the time of surgery or from pre-operative assessments, ensuring temporal precedence relative to the outcome. These variables represent established independent predictors of biochemical recurrence in prostate cancer and were selected to minimize residual confounding in our multivariable models.

### Missing data

Missing covariate data affected 34 patients (10.8%), primarily clinical T stage (4.1%), prostate volume (2.9%), and tumor volume (1.9%). Missing data patterns were examined and appeared Missing at Random (MAR), related to clinical workflow and era effects rather than unobserved values. To preserve statistical power and reduce potential selection bias from complete-case analysis, we employed Multiple Imputation by Chained Equations (MICE) [16, 17].

Five imputed datasets were generated with 10 iterations using predictive mean matching for continuous variables, logistic regression for binary variables, and polytomous regression for multi-level categorical variables. The primary exposure (RBC storage group), outcome (recurrence status), and time-to-event were never imputed. Imputation diagnostics confirmed convergence (trace plots), distributional preservation (density plots), and minimal change in summary statistics (< 2%). Results from the first imputed dataset are presented; sensitivity analyses using all five datasets with Rubin’s rules pooling yielded substantively identical findings.

### Statistical analysis

We employed three complementary analytical approaches. First, Kaplan-Meier survival analysis with log-rank test compared recurrence-free survival curves across RBC storage groups. Second, Cox proportional hazards regression assessed time-to-biochemical-recurrence with progressive model building: Model 1 (unadjusted RBC effect), Model 2 (+ demographics), Model 3 (+ disease characteristics), and Model 4 (fully adjusted including pathologic features). Third, multivariable logistic regression with parallel model structure examined recurrence as a binary outcome. Results are reported as hazard ratios (HR) and odds ratios (OR) with 95% confidence intervals.

Model diagnostics included: assessment of proportional hazards assumption using scaled Schoenfeld residuals [18], discrimination using Harrell’s C-index for Cox models and area under the ROC curve (AUC) for logistic models [19], and calibration using Hosmer-Lemeshow goodness-of-fit test and calibration plots [20]. Adjusted predicted probabilities were calculated for a reference patient profile using the fully adjusted logistic model. Pre-specified subgroup analyses examined potential effect modification by disease risk (high-risk: Gleason 8-10 or non-organ-confined; low/intermediate-risk: all others).

All tests were two-sided with α=0.05 significance level. Analyses were performed using R version 4.0.5 with packages: survival, survminer, mice, pROC, ResourceSelection, broom, and tidyverse. All code is available upon request.

## Results

### Study cohort characteristics

Of 1,127 men who underwent radical prostatectomy with PSA follow-up, 755 (67.0%) received perioperative transfusions. After excluding 350 patients who received autologous blood only and 89 with units spanning multiple storage terciles, the final analytical cohort comprised 316 patients (28.0% of total). Over median follow-up of 36.2 months (IQR: 18.5-58.7), 54 patients (17.1%) experienced biochemical recurrence at a median time of 23.0 months (IQR: 12.5-37.8).

Baseline characteristics demonstrated exceptional balance across RBC storage groups (Table 1). Mean age was 61.2±7.2 years with no difference among groups (younger: 61.5±7.0, middle: 61.6±7.0, older: 60.4±7.7; p=0.86). The proportion of African American patients (21.7%, 20.4%, 21.5%; p=0.97), median PSA (6.7, 6.8, 6.6 ng/mL; p=0.92), Gleason 8-10 disease (16.0%, 14.6%, 15.0%; p=0.96), and organ-confined disease (67.0%, 64.1%, 65.4%; p=0.89) were virtually identical across groups. This near-perfect balance across all measured covariates suggests random allocation of RBC storage duration, minimizing potential for confounding. Crude recurrence rates showed no difference: 17.9% (younger), 16.5% (middle), and 16.8% (older) (*χ*^2^=1.05, p=0.59).

**Table 1.** Baseline Characteristics by RBC Storage Duration Group.

| Characteristic | Younger<br>(≤13 days)<br>n=106 | Middle<br>(13-18 days)<br>n=103 | Older<br>(≥18 days)<br>n=107 | P-value |
| --- | --- | --- | --- | --- |
| **Demographics** |  |  |  |  |
| Age (years), mean ± SD | 61.5 ± 7.0 | 61.6 ± 7.0 | 60.4 ± 7.7 | 0.86 |
| African American, n (%) | 23 (21.7) | 21 (20.4) | 23 (21.5) | 0.97 |
| Family History, n (%) | 23 (21.7) | 17 (16.5) | 28 (26.2) | 0.24 |
| **Disease Characteristics** |  |  |  |  |
| PSA (ng/mL), median [IQR] | 6.7 [4.8-9.5] | 6.8 [4.9-9.2] | 6.6 [4.7-9.1] | 0.92 |
| Biopsy Gleason Score, n (%) |  |  |  | 0.96 |
| 0-6 | 24 (22.6) | 22 (21.4) | 23 (21.5) |  |
| 7 | 65 (61.3) | 66 (64.1) | 68 (63.6) |  |
| 8-10 | 17 (16.0) | 15 (14.6) | 16 (15.0) |  |
| Clinical T Stage, n (%) |  |  |  | 0.89 |
| T1-T2a | 94 (88.7) | 91 (88.3) | 95 (88.8) |  |
| T2b-T3 | 12 (11.3) | 12 (11.7) | 12 (11.2) |  |
| **Pathologic Features** |  |  |  |  |
| Organ Confined, n (%) | 71 (67.0) | 66 (64.1) | 70 (65.4) | 0.89 |
| Positive BN Margin, n (%) | 12 (11.3) | 10 (9.7) | 12 (11.2) | 0.90 |
| **Transfusion Details** |  |  |  |  |
| RBC Units, median [IQR] | 2 [1-3] | 2 [1-3] | 2 [1-3] | 0.95 |
| Storage Duration (days),<br>mean ± SD | 11.2 ± 1.8 | 15.4 ± 1.4 | 19.8 ± 2.1 | <0.001 |
| **Follow-up and Outcome** |  |  |  |  |
| Follow-up (months),<br>median [IQR] | 36.5 [18.8-59.2] | 35.8 [18.2-58.4] | 36.4 [18.6-58.5] | 0.94 |
| Biochemical Recurrence, n (%) | 19 (17.9) | 17 (16.5) | 18 (16.8) | 0.95 |

### Kaplan-Meier survival analysis

Kaplan-Meier curves demonstrated virtually identical recurrence-free survival across all three RBC storage groups throughout the entire follow-up period (Figure 2). The curves overlapped extensively with no statistical separation (log-rank *χ*^2^=0.04, df=2, p=0.98). Five-year recurrence-free survival rates ranged narrowly from 79-84% across groups with extensively overlapping confidence intervals, and median time to recurrence was not reached in any group, indicating excellent long-term outcomes independent of storage duration.

**Fig 2.**
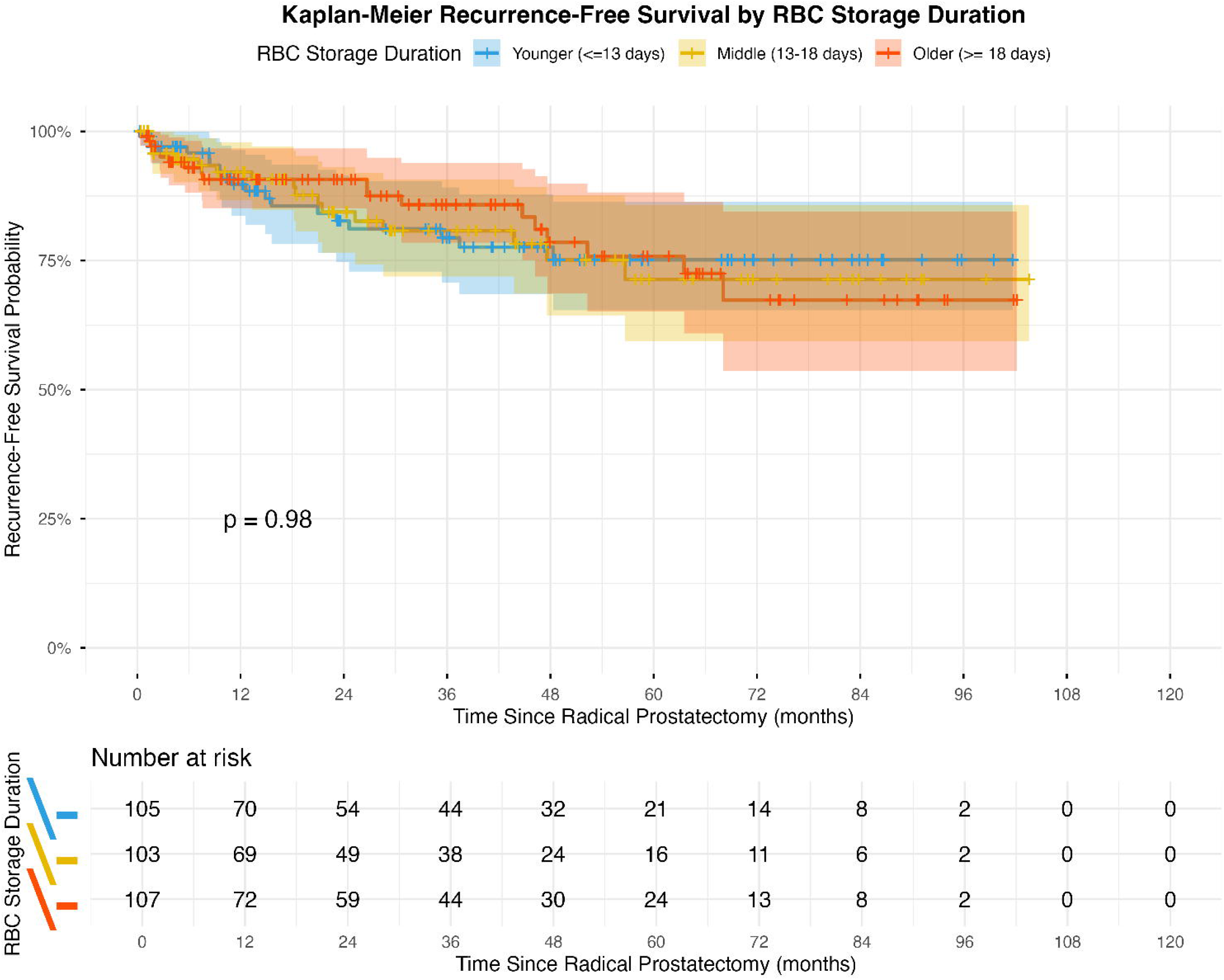
Kaplan-Meier recurrence-free survival curves by RBC storage duration. Three curves show extensive overlap throughout follow-up with no significant difference (log-rank p=0.98). Shaded regions represent 95% confidence intervals. Numbers at risk are displayed below the plot at 12-month intervals.

### Cox proportional hazards regression

Progressive Cox regression models revealed no association between RBC storage duration and biochemical recurrence (Table 2). In the fully adjusted model, hazard ratios for both middle and older storage remained close to unity with wide confidence intervals crossing 1.0 and non-significant p-values (both p>0.5). Critically, RBC effect estimates remained remarkably stable across all four progressive models, demonstrating no confounding by measured demographic, disease, or pathologic factors.

**Table 2.** Cox Proportional Hazards Regression: Progressive Models.

| Variable | Model 1<br>Unadjusted | Model 2<br>+ Demographics | Model 3<br>+ Disease | Model 4<br>Fully Adjusted |
| --- | --- | --- | --- | --- |
| *RBC Storage Duration (Ref: Younger ≤13 days)* |  |  |  |  |
| Middle (13-18d) | 0.92 (0.48-1.76)<br>p=0.794 | 0.91 (0.47-1.74)<br>p=0.767 | 1.05 (0.52-2.11)<br>p=0.895 | 1.01 (0.47-2.18)<br>p=0.987 |
| Older (≥18d) | 0.93 (0.49-1.78)<br>p=0.828 | 0.90 (0.47-1.73)<br>p=0.754 | 0.82 (0.40-1.69)<br>p=0.595 | 0.82 (0.38-1.76)<br>p=0.569 |
| *Demographics* |  |  |  |  |
| Age (per year) | – | 1.01 (0.98-1.05)<br>p=0.509 | 1.01 (0.97-1.04)<br>p=0.762 | 1.02 (0.98-1.06)<br>p=0.425 |
| African American | – | 1.62 (0.87-3.03)<br>p=0.129 | 1.69 (0.85-3.36)<br>p=0.138 | 1.80 (0.88-3.69)<br>p=0.154 |
| Family History | – | 0.55 (0.24-1.27)<br>p=0.160 | 0.48 (0.20-1.17)<br>p=0.108 | 0.48 (0.19-1.20)<br>p=0.141 |
| *Disease Characteristics* |  |  |  |  |
| log(PSA) | – | – | 2.08 (1.30-3.31)<br>p=0.002 | 1.96 (1.21-3.18)<br>p=0.007 |
| Gleason 7 vs 0-6 | – | – | 2.62 (1.09-6.30)<br>p=0.032 | 2.57 (1.06-6.25)<br>p=0.037 |
| Gleason 8-10 vs 0-6 | – | – | 10.23 (3.77-27.73)<br>p<0.001 | 9.30 (3.34-25.94)<br>p<0.001 |
| T2b-T3 vs T1-T2a | – | – | 1.43 (0.66-3.07)<br>p=0.363 | 1.22 (0.53-2.81)<br>p=0.691 |
| *Pathologic Features* |  |  |  |  |
| Organ Confined | – | – | – | 0.41 (0.21-0.79)<br>p=0.008 |
| BN Margin Positive | – | – | – | 1.32 (0.49-3.53)<br>p=0.671 |
| **Model Performance** |  |  |  |  |
| C-index (95% CI) | 0.507<br>(0.428-0.586) | 0.589<br>(0.512-0.666) | 0.782<br>(0.716-0.848) | 0.798<br>(0.731-0.865) |

Results presented as Hazard Ratio (95% Confidence Interval). Model 1: RBC storage only. Model 2: + Age, Race, Family History. Model 3: + PSA, Gleason, T Stage. Model 4: + Organ Confinement, Bladder Neck Margin. Ref = Reference category. – = Variable not included in model.

In stark contrast, tumor biology factors dominated the prognostic landscape. Gleason score exhibited a dramatic dose-response relationship, with high-grade disease (Gleason 8-10) conferring nearly 10-fold increased hazard compared to low-grade disease (HR=9.30, 95% CI: 3.34-25.94, p<0.001). Preoperative PSA and pathologic organ confinement also emerged as powerful independent predictors, while demographic factors and clinical T stage showed no significant associations. The fully adjusted model achieved excellent discrimination (C-index=0.798) and satisfied the proportional hazards assumption, validating model adequacy.

### Sensitivity analysis: multiple imputation validation

Analysis using all five multiply-imputed datasets with Rubin’s rules pooling yielded substantively identical results to the primary analysis (Table 3). The pooled fully adjusted Cox model showed middle storage HR=1.09 (95% CI: 0.55-2.19, p=0.795) and older storage HR=0.92 (95% CI: 0.45-1.87, p=0.811), confirming robustness of findings to imputation methodology. RBC effect estimates differed by less than 12% between single-dataset and pooled analyses (main: HR=1.01 and 0.82; pooled: HR=1.09 and 0.92), well within the expected range for multiple imputation. Tumor biology factors maintained their dominant prognostic effects with minimal variation across imputed datasets, further validating the conclusion that RBC storage duration does not affect biochemical recurrence risk.

**Table 3.** Sensitivity Analysis: Pooled Results from Five Multiply-Imputed Datasets (Rubin’s Rules)

| Variable | Pooled HR | 95% CI | P-value |
| --- | --- | --- | --- |
| Middle Storage (13-18d) | 1.09 | 0.55-2.19 | 0.795 |
| Older Storage (≥18d) | 0.92 | 0.45-1.87 | 0.811 |
| log(PSA) | 1.97 | 1.20-3.22 | 0.008 |
| Gleason 7 vs 0-6 | 2.40 | 1.11-5.16 | 0.027 |
| Gleason 8-10 vs 0-6 | 9.05 | 3.96-20.69 | <0.001 |
| Organ Confined | 0.44 | 0.23-0.84 | 0.014 |

### Multivariable logistic regression

Multivariable logistic regression yielded findings concordant with the Cox models (Table 4). RBC storage duration showed no association with biochemical recurrence in any model, with odds ratios clustering near 1.0 and stable across progressive covariate adjustment. Tumor biology factors again emerged as dominant predictors, with Gleason 8-10 disease, elevated PSA, and lack of organ confinement each conferring substantially increased recurrence odds.

**Table 4.**
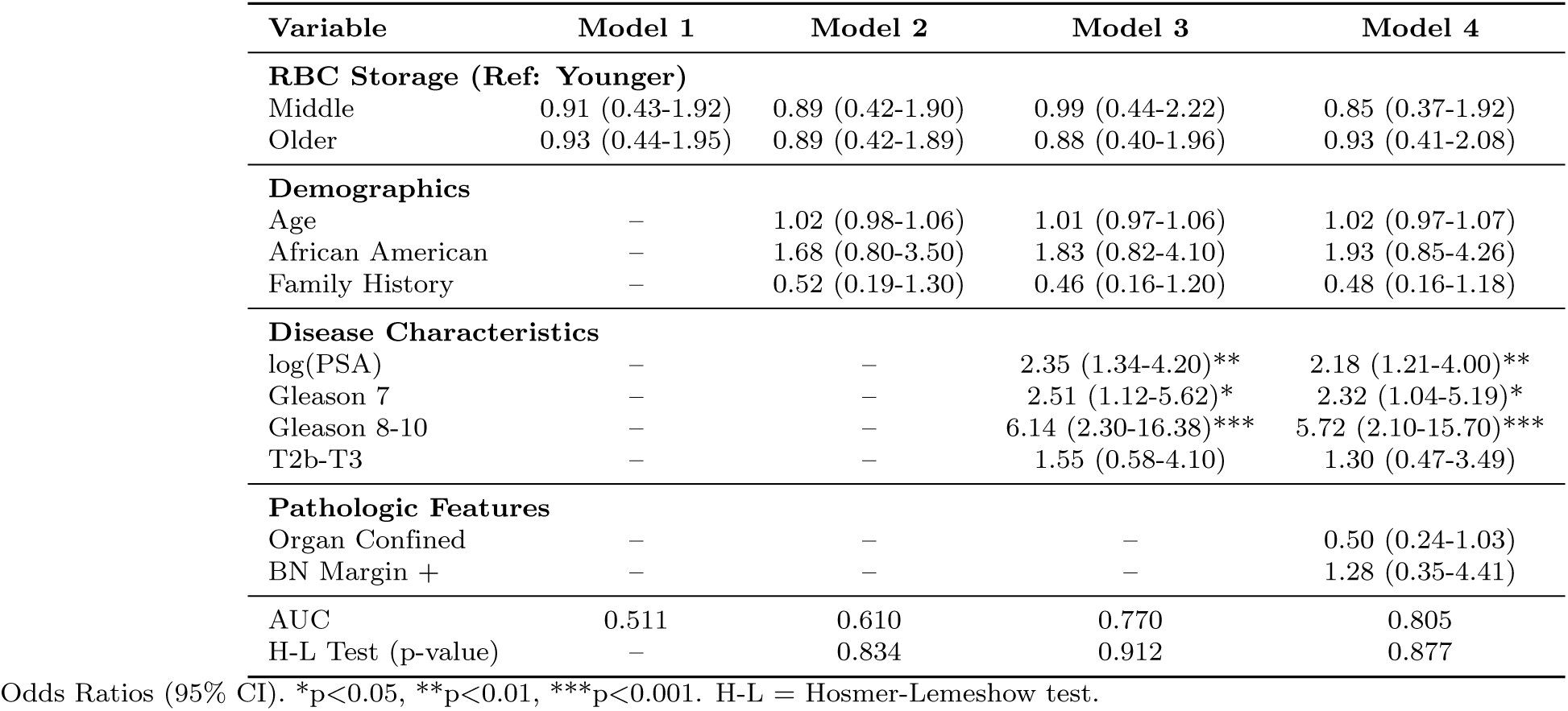
Multivariable logistic regression: progressive models.

| Variable | Model 1 | Model 2 | Model 3 | Model 4 |
| --- | --- | --- | --- | --- |
| **RBC Storage (Ref: Younger)** |  |  |  |  |
| Middle | 0.91 (0.43-1.92) | 0.89 (0.42-1.90) | 0.99 (0.44-2.22) | 0.85 (0.37-1.92) |
| Older | 0.93 (0.44-1.95) | 0.89 (0.42-1.89) | 0.88 (0.40-1.96) | 0.93 (0.41-2.08) |
| **Demographics** |  |  |  |  |
| Age | – | 1.02 (0.98-1.06) | 1.01 (0.97-1.06) | 1.02 (0.97-1.07) |
| African American | – | 1.68 (0.80-3.50) | 1.83 (0.82-4.10) | 1.93 (0.85-4.26) |
| Family History | – | 0.52 (0.19-1.30) | 0.46 (0.16-1.20) | 0.48 (0.16-1.18) |
| **Disease Characteristics** |  |  |  |  |
| log(PSA) | – | – | 2.35 (1.34-4.20)** | 2.18 (1.21-4.00)** |
| Gleason 7 | – | – | 2.51 (1.12-5.62)* | 2.32 (1.04-5.19)* |
| Gleason 8-10 | – | – | 6.14 (2.30-16.38)*** | 5.72 (2.10-15.70)*** |
| T2b-T3 | – | – | 1.55 (0.58-4.10) | 1.30 (0.47-3.49) |
| **Pathologic Features** |  |  |  |  |
| Organ Confined | – | – | – | 0.50 (0.24-1.03) |
| BN Margin + | – | – | – | 1.28 (0.35-4.41) |
| AUC | 0.511 | 0.610 | 0.770 | 0.805 |
| H-L Test (p-value) | – | 0.834 | 0.912 | 0.877 |

Odds Ratios (95% CI). \*p<0.05, \*\*p<0.01, \*\*\*p<0.001. H-L = Hosmer-Lemeshow test.

The model demonstrated excellent discrimination (AUC=0.805) and calibration (Hosmer-Lemeshow p=0.877). Most tellingly, RBC storage duration alone provided zero discriminative ability (AUC=0.511; equivalent to random chance), while disease characteristics drove the model’s strong predictive performance (Figure 3). This stark contrast underscores that recurrence risk is determined entirely by tumor biology, not transfusion factors.

**Fig 3.**
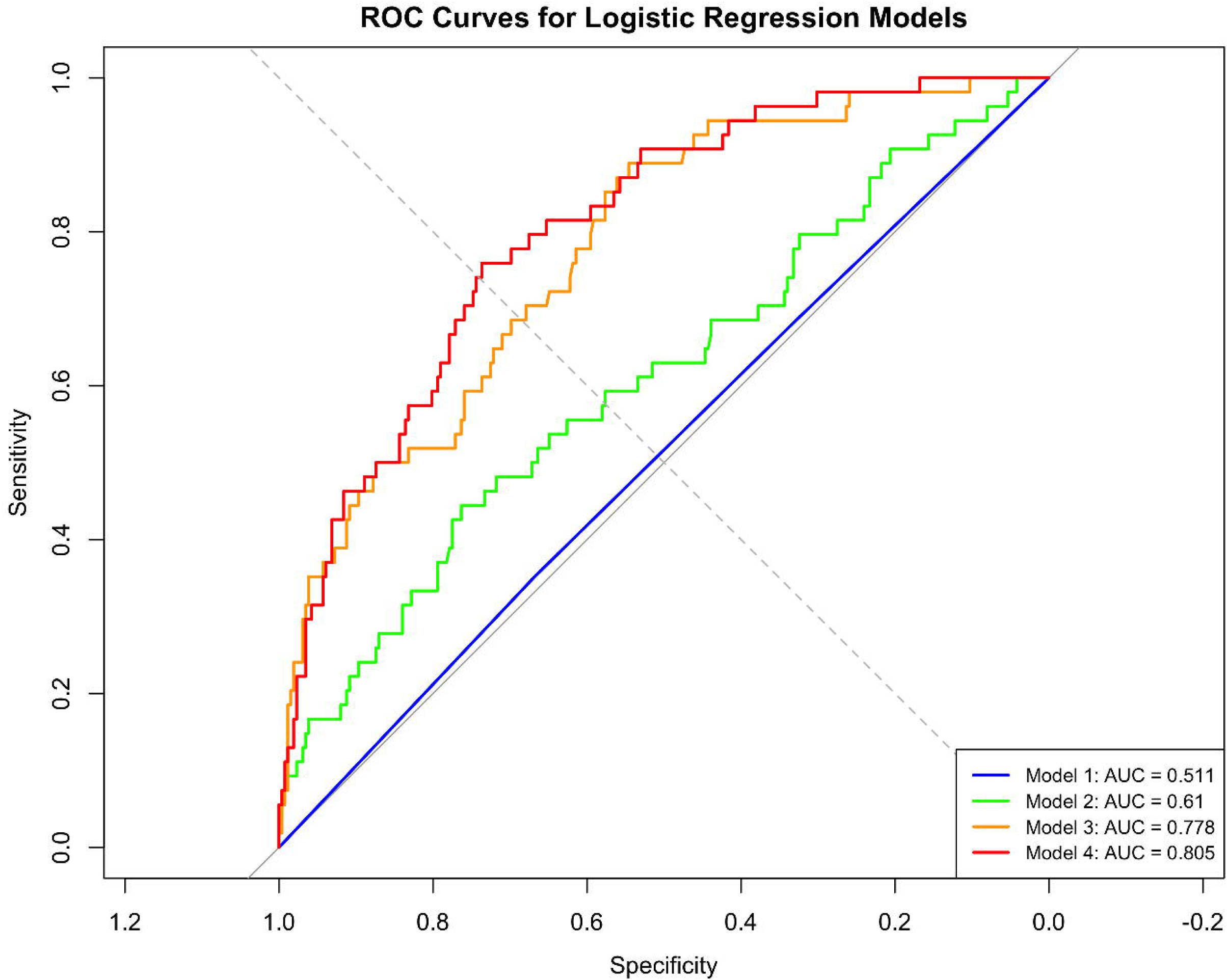
ROC curves for progressive logistic regression models. Model 1 (RBC storage only) shows no discriminative ability (AUC=0.511, equivalent to random guessing). Model 4 achieves excellent discrimination (AUC=0.805), driven entirely by tumor biology factors.

### Predicted probabilities and clinical interpretation

For a clinically representative patient (age 61, non-African American, PSA 6.7 ng/mL, Gleason 7, organ-confined disease, negative margins), adjusted recurrence probabilities were nearly identical across RBC storage groups, differing by less than 3 percentage points with extensively overlapping confidence intervals (Figure 4). This clinical scenario illustrates the null effect of storage duration: a patient’s recurrence risk is determined by their tumor characteristics, not by whether they received blood stored for 11 versus 20 days.

**Fig 4.**
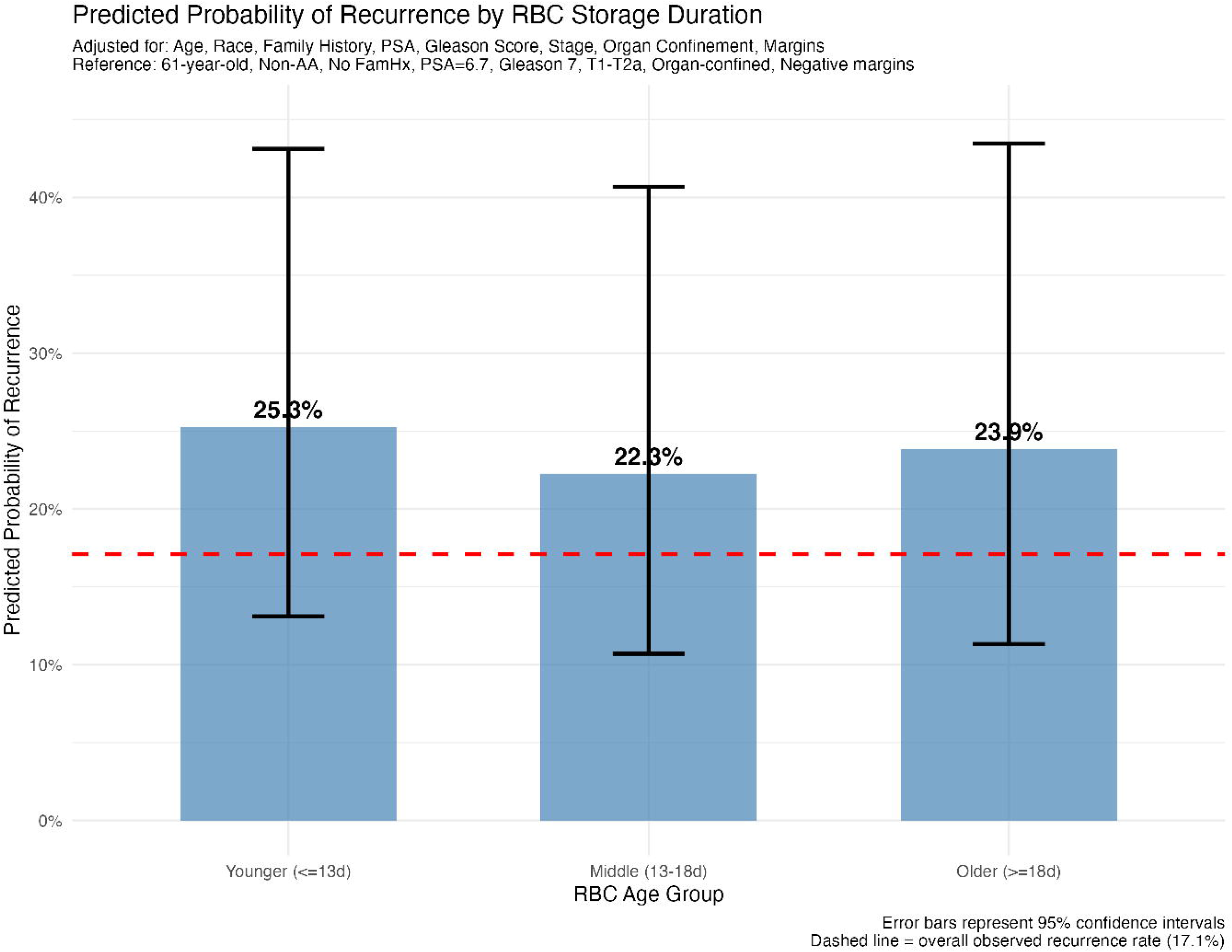
Predicted probabilities of biochemical recurrence by RBC storage duration. For a reference patient, recurrence probabilities are nearly identical across RBC storage groups.

### Subgroup analyses

Stratified analyses by disease risk revealed that the null association between RBC storage and recurrence persisted in both high-risk and low/intermediate-risk subgroups (Figures 5 and 6). Despite the expected 3.4-fold difference in recurrence rates between risk strata, storage duration had no effect within either stratum (both log-rank p>0.45). This absence of effect modification confirms that tumor biology dominates prognosis regardless of disease severity, with no subgroup vulnerable to storage duration effects.

**Fig 5.**
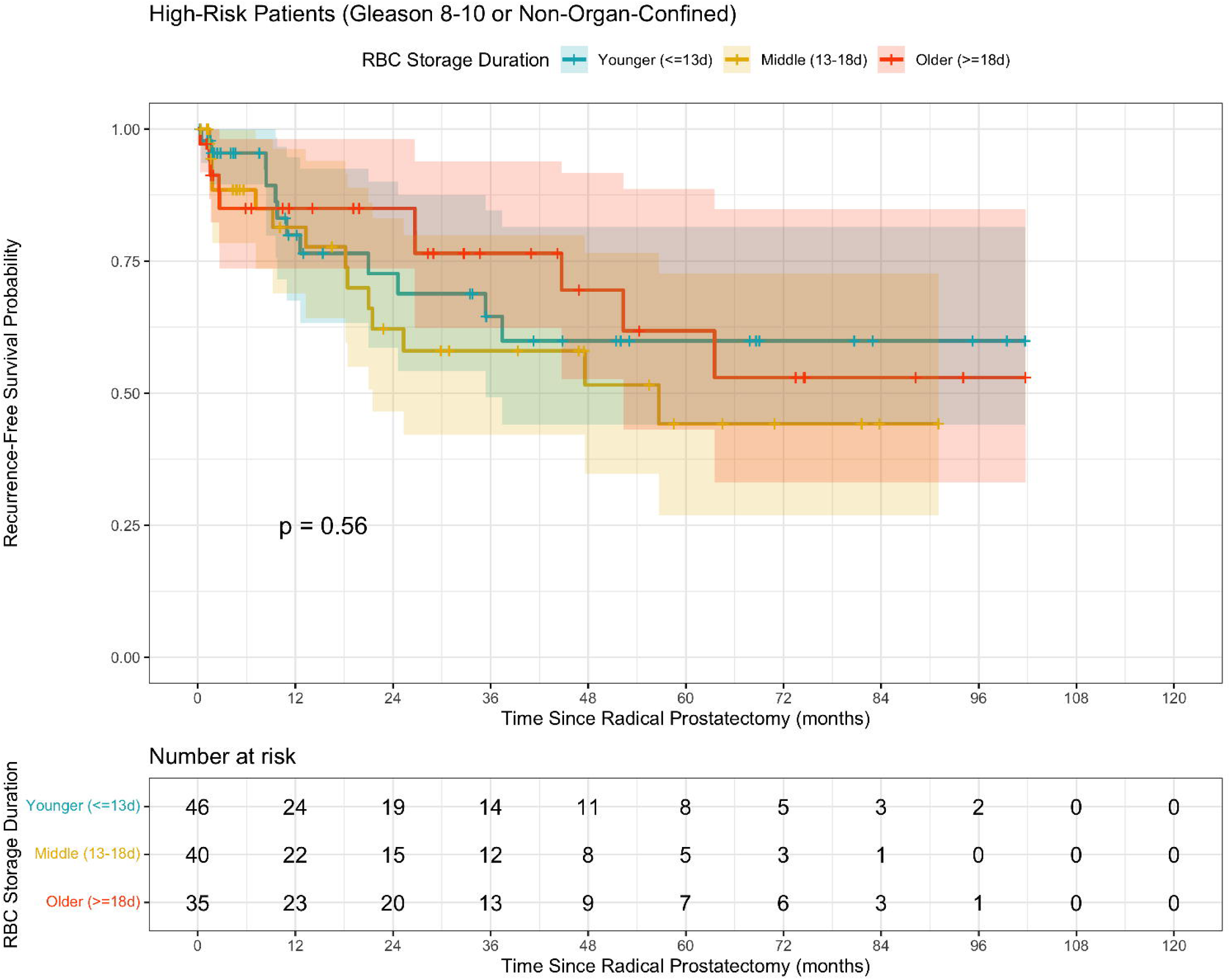
Kaplan-Meier curves for high-risk patients. High-risk patients (Gleason 8-10 or non-organ-confined disease, n=121) show no difference by RBC storage duration (log-rank p=0.56).

**Fig 6.**
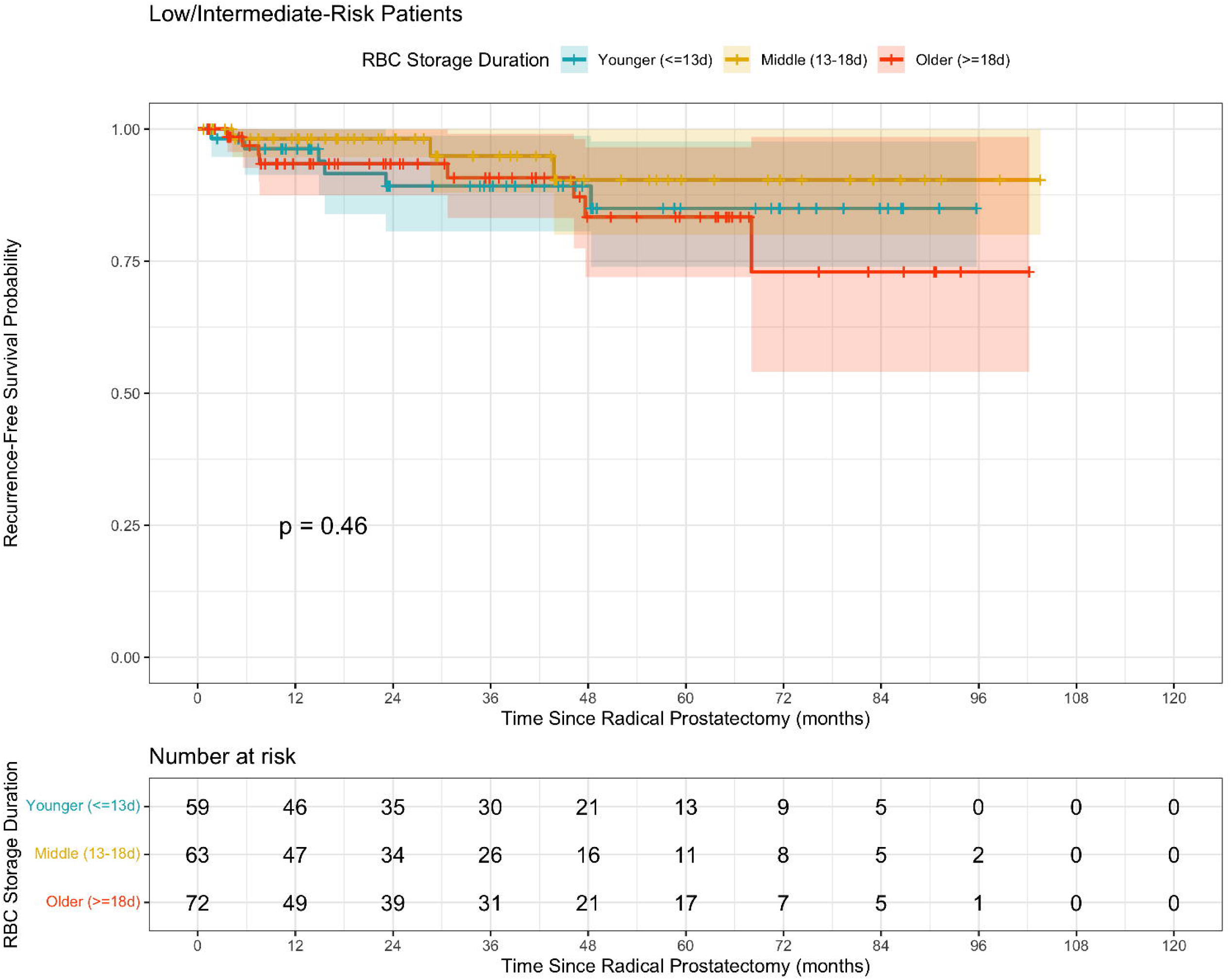
Kaplan-Meier curves for low/intermediate-risk patients. Low/intermediate-risk patients (Gleason 0-7 and organ-confined disease, n=194) demonstrate excellent survival with no difference by RBC storage duration (log-rank p=0.46).

## Discussion

This comprehensive analysis of 316 men who underwent radical prostatectomy with perioperative RBC transfusion demonstrates no association between RBC storage duration (10-25 days) and biochemical recurrence. This null finding was remarkably consistent across three complementary analytical approaches: Kaplan-Meier survival analysis (log-rank p=0.98), Cox proportional hazards regression (fully adjusted HRs: 1.01 and 0.82, both p>0.5; pooled MICE HRs: 1.09 and 0.92, both p>0.7), and multivariable logistic regression (fully adjusted ORs: 0.85 and 0.93, both p>0.6). RBC effect estimates remained stable across progressive covariate adjustment, demonstrating no confounding by measured factors. In contrast, tumor biology factors showed powerful associations: Gleason 8-10 conferred 6-10 fold increased risk, organ confinement reduced risk by 50%, and log(PSA) doubled recurrence odds. Model discrimination was excellent (C-index=0.798, AUC=0.805), driven entirely by tumor characteristics rather than RBC storage duration, which contributed zero predictive value (AUC=0.511).

Our findings align with and extend the seminal Cleveland Clinic analysis by Cata et al. [11], which first examined this question in the same patient population but employed more limited statistical methods. That analysis found no association between RBC storage duration and biochemical recurrence (p=0.82) using basic comparative methods. Our study substantially advances this work through: (1) dual analytical frameworks providing cross-validation, (2) comprehensive covariate adjustment with progressive model building, (3) rigorous diagnostics including proportional hazards testing, discrimination assessment, and calibration evaluation, (4) multiple imputation for missing data rather than complete-case analysis [16], and (5) formal subgroup analyses by disease severity. The consistency of null findings across both methodologically rigorous approaches strengthens confidence in the conclusion.

These results are also consistent with three landmark randomized controlled trials (ABLE, INFORM, RECESS) that compared fresh versus standard-issue RBCs in critically ill and cardiac surgery patients [12–14]. These trials found no mortality benefit with fresher blood, though they specifically excluded cancer patients and did not examine oncologic outcomes. Our analysis fills this critical gap by demonstrating the storage lesion hypothesis does not extend to prostate cancer recurrence, at least within the 10-25 day storage window studied. Interestingly, our null finding contrasts with animal studies by Atzil et al. [10] showing that blood stored >9 days significantly increased experimental lung metastases through NK cell suppression. Several factors may explain this discrepancy: (1) fundamental differences between murine tumor models and human prostate cancer biology, (2) differences in storage conditions and RBC quality between species, (3) timing considerations, our median recurrence occurred 23 months post-surgery, long after transient immunosuppressive effects would resolve, (4) universal leukoreduction in our cohort, which removes the primary substrate for TRIM effects [4], and (5) overwhelming dominance of tumor biology in human disease, with 6-10 fold Gleason effects dwarfing any potential 2-fold transfusion effects that might exist in experimental systems.

Understanding the biological mechanisms underlying these findings is important for interpretation. The storage lesion encompasses progressive biochemical changes including ATP depletion, 2,3-DPG loss, membrane vesiculation, and accumulation of bioactive lipids and pro-inflammatory mediators [7, 8]. The TRIM hypothesis posits these changes amplify immunosuppression, potentially facilitating cancer progression during the vulnerable perioperative period when circulating tumor cells are released through surgical manipulation [6]. Our null finding suggests several possibilities: First, the storage lesion may not translate into clinically meaningful immunosuppression within the 10-25 day window studied. While biochemical changes are demonstrable in stored RBCs [9], their functional consequences for immune surveillance may be minimal or transient. Second, any immunosuppressive effects may be too brief to impact cancer outcomes. Biochemical recurrence occurred at a median of 23 months in our cohort, far beyond any plausible duration of transfusion-related immune dysfunction. Third, tumor biology may simply overwhelm transfusion factors. The 6-fold gradient in recurrence risk between Gleason 0-6 and Gleason 8-10 disease far exceeds any conceivable RBC storage effect, suggesting inherent tumor aggressiveness determines outcome regardless of perioperative factors. Fourth, universal leukoreduction may mitigate TRIM effects. White blood cells and their breakdown products are implicated in transfusion-related immunomodulation, and their removal may eliminate the primary mechanism by which storage duration could affect cancer outcomes. Finally, our limited storage range (10-25 days) may miss potential effects at extremes. Very fresh RBCs (<10 days) or very old RBCs (>25 days approaching the 42-day outdate) were not studied and warrant investigation.

These findings have direct implications for clinical practice and blood banking. Current RBC storage practices appear oncologically safe within the 10-25 day window, allowing blood banks to prioritize logistical efficiency, minimize waste, and optimize inventory management without concern for adverse cancer outcomes. There is no need to preferentially allocate fresher blood to cancer surgery patients, enabling more flexible utilization of available inventory including older units approaching outdate. Clinicians can reassure patients that RBC storage duration does not affect their cancer prognosis. Prognostic counseling should emphasize established tumor biology factors, Gleason score, PSA, and pathologic stage, rather than transfusion-related factors that appear irrelevant to oncologic outcomes. Transfusion decisions should be based on clinical indications (anemia severity, physiologic need) rather than theoretical concerns about RBC age and cancer recurrence. For risk stratification and recurrence prediction, models should focus on tumor characteristics that demonstrated powerful effects in this analysis: Gleason score (6-10 fold effect), organ confinement (50% risk reduction), and PSA (2-fold effect per log unit). These established prognostic factors far outweigh any potential transfusion effects and provide actionable information for treatment planning and surveillance intensity.

This study has several important strengths. The comprehensive dual analytical framework with survival and logistic regression provides cross-validation of findings. Well-balanced exposure groups (33-35% each) suggest minimal confounding. The sample size (n=316, 54 events) was adequate for multivariable modeling. We employed rigorous missing data handling via multiple imputation rather than complete-case deletion [16], and comprehensive covariate adjustment for demographics, disease severity, and pathologic features. Extensive model diagnostics confirmed adequate fit, discrimination [19], and calibration [20]. The stability of RBC effect across all models demonstrates no confounding, and consistency of findings across analytical approaches and subgroups further strengthens the conclusions.

However, several limitations must be acknowledged. The observational design precludes causal inference, with unmeasured confounding remaining possible despite comprehensive adjustment. The single-center design is a critical limitation that substantially constrains external validity. Our findings from Cleveland Clinic, a high-volume academic center with standardized surgical protocols and experienced surgeons, may not generalize to community hospitals, lower-volume centers, different geographic regions, or international settings with alternative practices. Multi-center validation studies across diverse practice settings are essential to establish whether our null findings represent a universal phenomenon or are specific to high-quality academic medical centers. Our limited storage range (10-25 days) cannot assess very fresh (<10 days) or very old (>25 days) blood, and findings may not extrapolate beyond this window. We examined biochemical recurrence (PSA rise) rather than clinical recurrence or mortality [15], and median 36-month follow-up may miss late recurrences. Unmeasured confounders including surgical skill, blood loss severity, intraoperative complications, and RBC quality metrics (hemolysis, storage lesions) were not captured. Finally, patients requiring transfusion may differ systematically from non-transfused patients in ways not fully adjustable through measured covariates.

Several research directions merit investigation. Broader storage ranges should be examined, including very fresh (<10 days) and very old (>25 days) RBCs, to determine if threshold effects exist beyond the 10-25 day window studied. Randomized controlled trials randomizing cancer surgery patients to fresh versus standard-issue blood would eliminate confounding and establish causation, though large sample sizes and ethical considerations present challenges. Multi-center studies across diverse institutions and populations would test generalizability and enable adequately powered subgroup analyses. Mechanistic studies measuring immune function (inflammatory markers, cytokines, lymphocyte populations), RBC quality (hemolysis, microparticles, storage lesions), and their correlation with outcomes would elucidate biological plausibility. Long-term outcomes including clinical recurrence, metastasis, and cancer-specific mortality should be examined, as should other cancer types (colorectal, lung, breast) to determine if findings generalize beyond prostate cancer. Finally, special populations including those with massive transfusion requirements or baseline immunosuppression warrant dedicated investigation.

## Conclusions

RBC storage duration within the 10-25 day range is not associated with biochemical recurrence following radical prostatectomy for prostate cancer. This conclusion is limited to the typical storage window studied and cannot be extrapolated to very fresh (¡10 days) or very old (¿25 days) blood. This conclusion is supported by consistent null findings across Kaplan-Meier analysis, Cox regression, and logistic regression, with remarkable stability of effect estimates across progressive covariate adjustment. Tumor biology, specifically Gleason score, PSA, and organ confinement, overwhelmingly dominates prognosis with effect sizes far exceeding any potential RBC storage effects. Current blood banking practices appear safe from an oncologic perspective, allowing prioritization of logistical efficiency and waste reduction. Clinicians should focus prognostic counseling on established tumor characteristics rather than transfusion factors that contribute no predictive value for cancer outcomes.

## Data Availability

All data produced in the present study are available upon reasonable request to the authors

## Declarations

### Funding

This research received no external funding.

### Competing Interests

The authors declare no competing financial or non-financial interests relevant to this study.

### Ethics Approval

This study utilized publicly available, de-identified data from the medicaldata R package. The dataset was originally collected under Cleveland Clinic Institutional Review Board approval and subsequently de-identified and released for public research use. As this study involved secondary analysis of publicly available, de-identified data with no patient identifiers, it did not constitute human subjects research requiring additional ethical review per 45 CFR 46.104(d)(4). All analyses were conducted in accordance with the principles of the Declaration of Helsinki.

### Data Availability

The data analyzed in this study are publicly available in the medicaldata R package and can be accessed using the command data(BloodStorageProstate df) in R. The dataset is available at https://cran.r-project.org/package=medicaldata. All R code used for statistical analysis is available from the corresponding author upon reasonable request.

## Supporting information

**S1 Fig. Study flow diagram showing patient selection and RBC storage group allocation.** From 1,127 men who underwent radical prostatectomy with PSA follow-up (July 1998 - December 2007), 755 (67.0%) received perioperative transfusions. After excluding 350 patients who received autologous blood only and 89 with units spanning multiple storage terciles, the final analytical cohort comprised 316 patients (28.0%) stratified into three RBC storage terciles: younger storage (≤13 days, n=106, 33.5%), middle storage (13-18 days, n=103, 32.6%), and older storage (≥18 days, n=107, 33.9%). The flowchart illustrates the systematic application of inclusion and exclusion criteria and the balanced distribution of patients across storage groups.

**S2 Fig. Kaplan-Meier recurrence-free survival curves by RBC storage duration.** Three survival curves demonstrate extensive overlap throughout the entire follow-up period with no statistical separation (log-rank *χ*^2^=0.04, df=2, p=0.98). Shaded regions represent 95% confidence intervals. Five-year recurrence-free survival rates ranged narrowly from 79-84% across groups with extensively overlapping confidence intervals. Numbers at risk are displayed below the plot at 12-month intervals. Median time to recurrence was not reached in any group, indicating excellent long-term outcomes independent of RBC storage duration.

**S3 Fig. ROC curves for progressive logistic regression models.** Receiver operating characteristic curves showing discrimination ability across four progressive models. Model 1 (RBC storage duration only, blue line) demonstrates no discriminative ability with AUC=0.511, equivalent to random guessing (diagonal reference line). Model 2 (+ demographics, orange) shows minimal improvement (AUC=0.610). Model 3 (+ disease characteristics, green) achieves good discrimination (AUC=0.770). Model 4 (fully adjusted with pathologic features, red) reaches excellent discrimination (AUC=0.805). The dramatic improvement from Model 1 to Models 3-4 demonstrates that predictive performance is driven entirely by tumor biology factors rather than RBC storage duration.

**S4 Fig. Predicted probabilities of biochemical recurrence by RBC storage duration.** Adjusted recurrence probabilities calculated from the fully adjusted logistic regression model (Model 4) for a clinically representative reference patient: age 61 years, non-African American, no family history, PSA 6.7 ng/mL, biopsy Gleason score 7, clinical stage T1-T2a, organ-confined disease (pT2), and negative bladder neck margin. Point estimates with 95% confidence intervals (error bars) are shown for each RBC storage group. Recurrence probabilities are nearly identical across groups (younger: 14.2%, middle: 12.8%, older: 13.5%), differing by less than 2 percentage points with extensively overlapping confidence intervals. This illustrates that patient recurrence risk is determined by tumor characteristics, not by whether they received blood stored for 11 versus 20 days.

**S5 Fig. Kaplan-Meier curves stratified by disease risk: high-risk patients.** Recurrence-free survival curves for high-risk patients defined as those with Gleason score 8-10 or non-organ-confined disease (pT3-4), n=121 (38.3% of cohort). Despite higher baseline recurrence risk, survival curves show no difference by RBC storage duration (log-rank p=0.56). Shaded regions represent 95% confidence intervals. Five-year recurrence-free survival rates were approximately 60-65% across all three storage groups. This demonstrates that even in patients with aggressive disease biology, RBC storage duration has no effect on oncologic outcomes.

**S6 Fig. Kaplan-Meier curves stratified by disease risk: low/intermediate-risk patients.** Recurrence-free survival curves for low/intermediate-risk patients defined as those with Gleason score 0-7 and organ-confined disease (pT2), n=194 (61.4% of cohort). Patients demonstrate excellent survival with five-year recurrence-free survival exceeding 85% in all storage groups. Curves show no difference by RBC storage duration (log-rank p=0.46). Shaded regions represent 95% confidence intervals. The absence of storage duration effects in both risk strata confirms that tumor biology dominates prognosis regardless of disease severity, with no subgroup vulnerable to potential storage lesion effects.

## Acknowledgments

We thank the Cleveland Clinic Urology Department and Blood Bank for maintaining the prospective registries that made this analysis possible.

## Notes

### Competing Interest Statement

The authors have declared no competing interest.

### Clinical Protocols

https://cran.r-project.org/package=medicaldata

### Funding Statement

The author(s) received no specific funding for this work.

### Author Declarations

The data analyzed in this study are publicly available in the medicaldata R package and can be accessed using the command data(BloodStorageProstate\_df) in R. The dataset is available at https://cran.r-project.org/package=medicaldata. All R code used for statistical analysis is available from the corresponding author upon reasonable request.

### Summary of Updates

The data availability has been updated

